# A comparative analysis of serial measurements of Soluble Urokinase-type Plasminogen Activator Receptor (suPAR) and C-reactive protein in patients with moderate COVID-19: a single center study from India

**DOI:** 10.1101/2022.03.30.22273026

**Authors:** Atul Kakar, Pooja Rani, Tanvi Batra, Rizwana Hasan, Sangeeta Choudhury

**Affiliations:** Department of Internal Medicine, Sir Ganga Ram Hospital, India; Department of Research, Sir Ganga Ram Hospital, New Delhi, India

**Keywords:** COVID-19, soluble urokinase-type plasminogen activator receptor, C-reactive protein, biomarkers, comorbid conditions, mortality

## Abstract

Soluble urokinase plasminogen-activator receptor (suPAR) is a secreted protein associated with inflammation and proven its usefulness in triage/risk stratifications. This prospective study aimed to evaluate the utility of suPAR in comparison to C-reactive protein (CRP) in hospitalized moderate COVID-19 patients.

This is a prospective comparative study during second pandemic wave. Serum suPAR level and CRP were measured serially in 31 confirmed COVID-19 hospitalized patients (20 males, 11 females) on day-1 (24-hours of admission), day-3 and day-5 using suPARnostic AUTO flex ELISA and Nephelometry (ThermoFischer) respectively. Schapiro Wilk test verified the data distribution; Wilcoxon signed rank test compared CRP and suPAR between deceased/alive subject and identified link between co-morbidity and COVID-19 severity.

In our study, the mean age was 61.8 ranging from 28-82 with 9.7% (n=3/31) mortality rate. Deceased patients showed significant higher suPAR levels correlating with increasing severity from day 1 to 5 (p<0.016-0.006) than CRP (p=0.717). Patients with pre-existing co-morbidities showed significantly elevated suPAR levels on days 1-5, especially those with hypertension (HTN;p<0.03) and chronic kidney disease (CKD;p<0.001).

In conclusion, levels of suPAR were higher in deceased patients with severe symptoms of COVID-19 during hospitalization and in patients with pre-existing co-morbid conditions, HTN and CKD. This preliminary study provides evidence suggesting that circulating suPAR can be a potential biomarker to assess the severity of COVID 19 compared to CRP.

## 1. Introduction

During the second wave of Novel Corona Virus disease 2019 (COVID-19), the delta variant had intensely affected on healthcare systems globally. In India, at the time of writ-ing this manuscript, there had been 40 million confirmed cases of COVID-19 with more than four lakh deaths (WHO COVID-19) [1]. In India, the cases started to increase around February 2021 and peaked between 15 March, 2021 to 30 June, 2021 as per the data depicted by the Worldometer [1]. On 01 March 2021, India reported daily 11,000 new cases with 80 deaths that steadily increased to 402,110 cases daily with 3,620 deaths on 30 April 2021 [1]. As on March 2022, the world records 440,947,092 cases with 5,995,271 death toll (India statistics - 42,945,160 total cases / 514,419 deaths). In the meanwhile, scientists have explored several options to develop preventive approaches like vaccines and treatment strategies ie., antiviral drugs (remdesivir, molnupiravir) including anti-inflammatory tocilizumab and monoclonal antibody cocktail. Yet, several individuals continue to die worldwide. Hence, a good biomarker is needed that can not only predict the disease sever-ity but can also offer information to the clinician if the disease status can lead to mortality.

Clinical studies observed that levels of blood markers like CRP (C-reactive protein) and erythrocyte sedimentation rate (ESR) were associated with increasing severity and mortality in subjects with COVID-19 [2,3,4]. CRP is synthesized primarily by hepatocytes [5] and is significantly associated with inflammatory processes [3,6]. It is an acute-phase protein reactant that increases following secretion of interleukin-6 (IL-6) by T-cells and macrophages under inflammatory conditions. Nevertheless, it offers limited prognostic information most likely because it binds to membrane protein lysophosphatidylcholine expressed by dead or dying cells [7]. Soluble urokinase-type plasminogen activator receptor (suPAR) is produced mainly by leukocytes and activated endothelium [8] under inflammatory condition and is found to be elevated in blood, plasma, serum, urine, bronchoalveolar lavage and cerebrospinal fluid [9]. This soluble form of urokinase-type plasminogen activator receptor (uPAR) is a glyosyl phosphatidylinositol (GPI) linked mem-brane protein of 55-60 kDa present in three forms (I-III, II-III and I) and is cleaved by proteases under inflammatory condition [10]. The receptor uPAR is present on various immunologically active cells including neutrophils, monocytes, macrophages, keratinocytes, smooth muscle cells, fibroblast, activated T-lymphocytes, endothelial cells [11].

In this prospective study, we serially measured suPAR and CRP, the two pro-inflammatory markers, in patients of moderate COVID-19 with the aim to compare their prognostic performance. Our data showed suPAR to be a better prognostic biomarker than CRP in moderate COVID-19 patients. Additionally suPAR was also identified as an independent risk factor for COVID-19 patient with pre-existing co-morbid factors.

## 2. Materials and Methods

### 2.1 Patient enrolment

This study was carried out during the second wave of COVID-19 (February to March 2021) at a tertiary care hospital in India. Only those patients who were admitted to hospital with COVID-19 positive were enrolled in the study. A total of 1007 were admitted during the second wave of infection. Fifty patients were admitted between February and March 2021, wherein 31 patients were enrolled as per the inclusion criteria [moderate COVID-19 as defined by Ministry of Health and Family Affair, India (MoH & FW); (Figure 1)]. Nineteen patients requiring noninvasive/ invasive mechanical ventilation and were excluded. Patients were followed up for 45 days from the time of admission to documented survival/mortality. History of co-morbid factors such as diabetes (DM), obesity, hyper-tension (HTN), chronic kidney disease (CKD), chronic obstructive airway disease (COPD) and coronary artery disease (CAD) was recorded. The study was reviewed and approved by Institutional Ethical Committee (# EC/08/20/1711)

**Figure 1.**
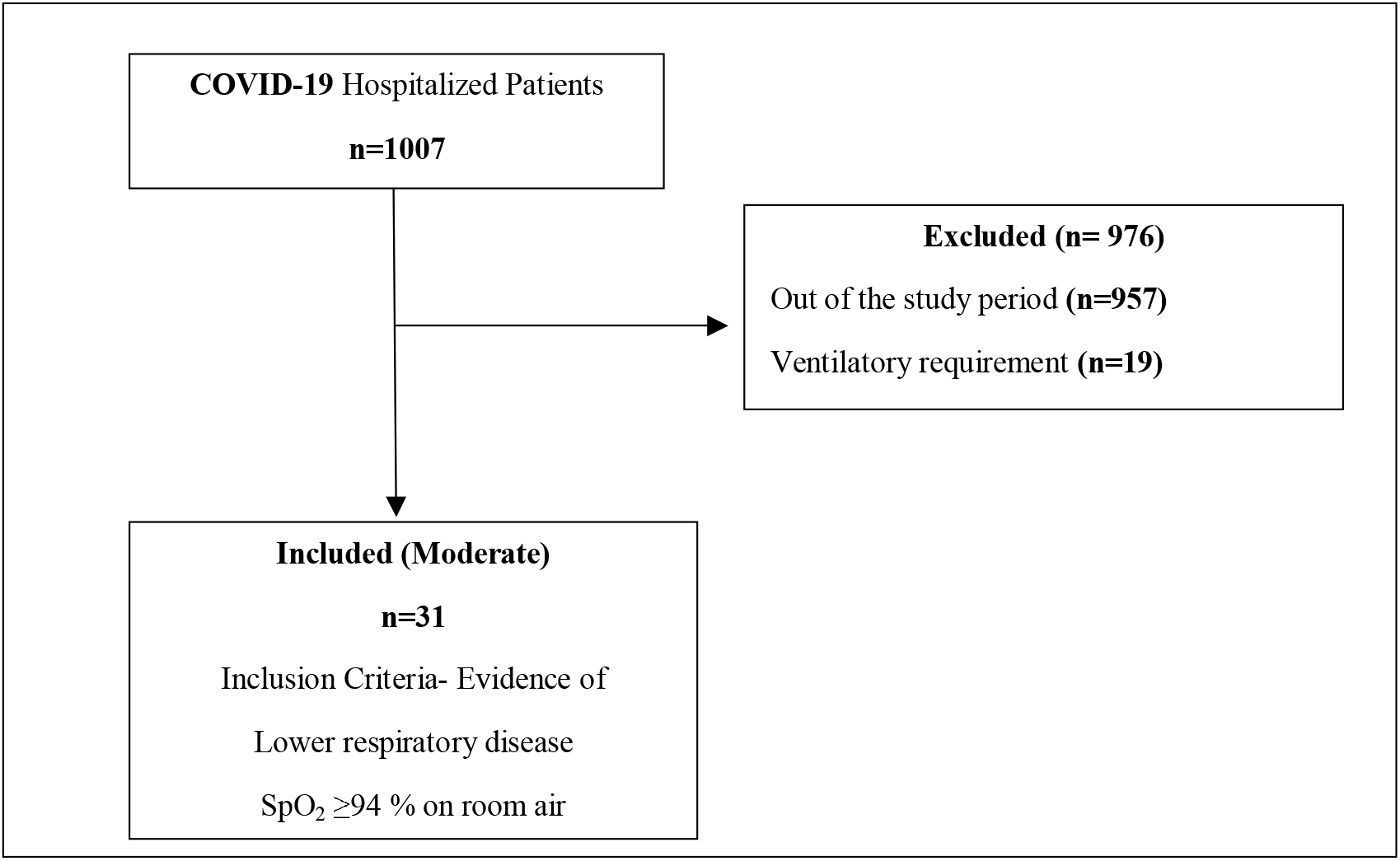
Flow chart of study sample and clinical definitions of mild, moderate and severe COVID-19 patients as defined (covid19 treatmentgudelines.nih.gov/overview/clinical-spectrum/)[12]

### 2.2 Assay protocol

Blood samples were collected on three time points i.e., day-1 of admission, day-3 and day-5 or on the day of discharge if discharged before 5 days. Blood sample of 2 ml was collected in non-EDTA/plain for biomarker (suPAR level) measurement. Sera was separated as per standardized protocol i.e., centrifuged at 2000g for 15 min and aliquots stored at -80°C until further use. The levels of suPAR were estimated using the suPARnostic AUTO flex® ELISA kit (ViroGates, Copenhagen, Denmark). The sensitivity of assay ranged from 0.6-20ng/ml as determined by the manufacturer. The normal range of suPAR in adults are 2-3ng/ml and levels above 9-10ng/ml is reported in critically ill patients. Nephelometry (Thermo Fischer) method was used to quantify CRP levels.

### 2.3 Statistical analysis

Statistical analysis was performed using the SPSS version 17.0 program for windows (SPSS Inc., Chicago, IL, USA). Data expressed as means (mean ± SEM). We conducted a Schapiro Wilk test to verify the distribution of the data. Wilcoxon signed rank test used for comparison of CRP and suPAR on Day-1, Day-3 and Day-5 in dead/alive. This test was also used to identify the link between co-morbidity and COVID-19 severity. P value of <0.05 was taken as significant.

## 3. Results

### 3.1 Demographics and clinical parameters

The study included 31 subjects who were hospitalized with moderate COVID-19 (confirmed RT-PCR for SARS-Cov-2). The mean ± SEM age of the moderate COVID-19 patients was 61.84 ± 2.17, ranging from 30-70 years. The ratio of male: female was 2:1.05. Out of all patients, 64.5% (20/31) were male aged 28-76 years and 35.5% were female aged 51-82 years. At the end of study, three (9.7%) hospitalization patients developed ARDS who later died. The demographics and the clinical presentation of the total cohort are shown in Table 1.

**Table 1:** General demographics and clinical presentations of patients.

| Characteristics | n (%) |
| --- | --- |
| **Age group** |  |
| >30 years | 3.2% (n=1/31) |
| 30-50 years | 6.5% (n=2/31) |
| 50-70 years | 61.3% (n=19/31) |
| >70 years | 29% (n=9/31) |
| **Mortality** | 9.7% (n=3/31) |
| **Length of Hospital stay (up to 45 days)** | 6 – 34 days |
| **Sex** |  |
| Male | 64.5% (n=20/31) |
| Female | 35.5% (n=11/31) |
| **Co-morbidities** |  |
| Hypertension | 54.8% (n=17/31) |
| Diabetes | 35.5 % (n=11/31) |
| Hypothyroid | 19.4% (n=6/31) |
| Chronic Kidney Disease | 12.9% (n=4/31) |
| COPD | 9.7%, (n=3/31) |
| CAD | 9.7% (n=3/31) |

Hypertension was the most common co-morbidity (17/31; 54.80%), followed by diabetes (11/31; 35.5%), hypothyroid (6/31; 19.4%), chronic kidney disease (4/31; 12.9%), COPD (3/31; 9.7%) and CAD (3/31; 9.7%) (Table 1)

### 3.2 Comparison between serum levels of suPAR and CRP

suPAR and CRP levels were measured in all enrolled moderate COVID-19 patients on the day of admission (day-1), day-3 and day-5 as shown in Table 2.

**Table 2.**
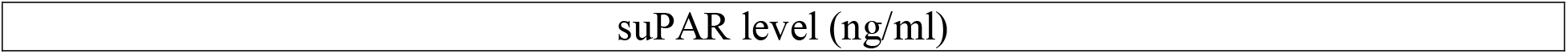
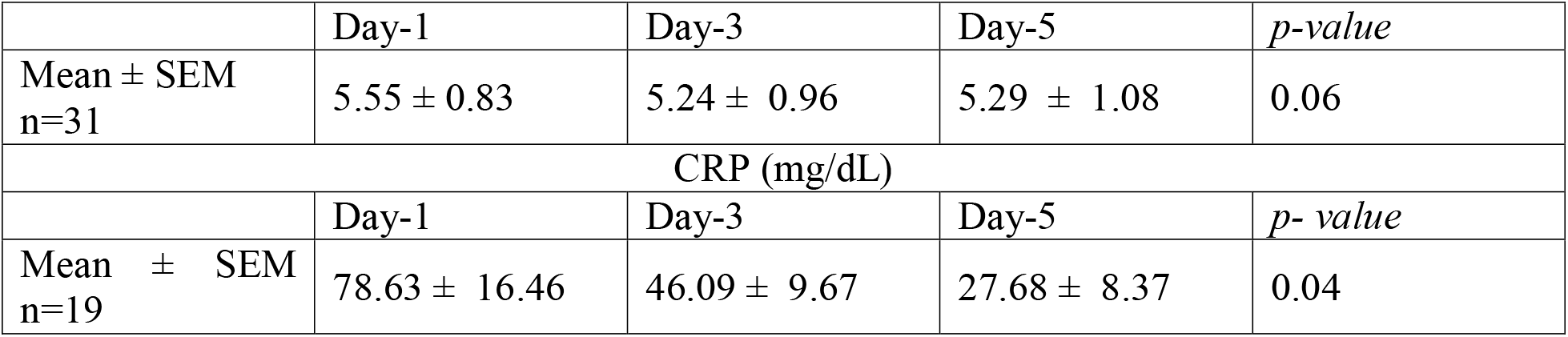
Level of suPAR versus CRP in COVID-19 patients on Days 1, 3 and 5.

| suPAR level (ng/ml) |
| --- |

|  | Day-1 | Day-3 | Day-5 | *p-value* |
| --- | --- | --- | --- | --- |
| Mean ± SEM<br>n=31 | 5.55 ± 0.83 | 5.24 ± 0.96 | 5.29 ± 1.08 | 0.06 |
| CRP (mg/dL) |  |  |  |  |
|  | Day-1 | Day-3 | Day-5 | *p-value* |
| Mean ± SEM<br>n=19 | 78.63 ± 16.46 | 46.09 ± 9.67 | 27.68 ± 8.37 | 0.04 |

A significant increased levels of serum suPAR was observed among deceased patients (n=3) on each day of evaluation - day-1 (15.28 ± 5.12); day-3 (17.3 ± 6.29); day-5 (19.6 ± 7.27) when compared to alive patient (n=28) on day-1 (4.51 ± 0.46); day-3 (3.95 ± 0.42); day-5 (3.7 ± 0.34) as shown in Figure 2.

**Figure 2:**
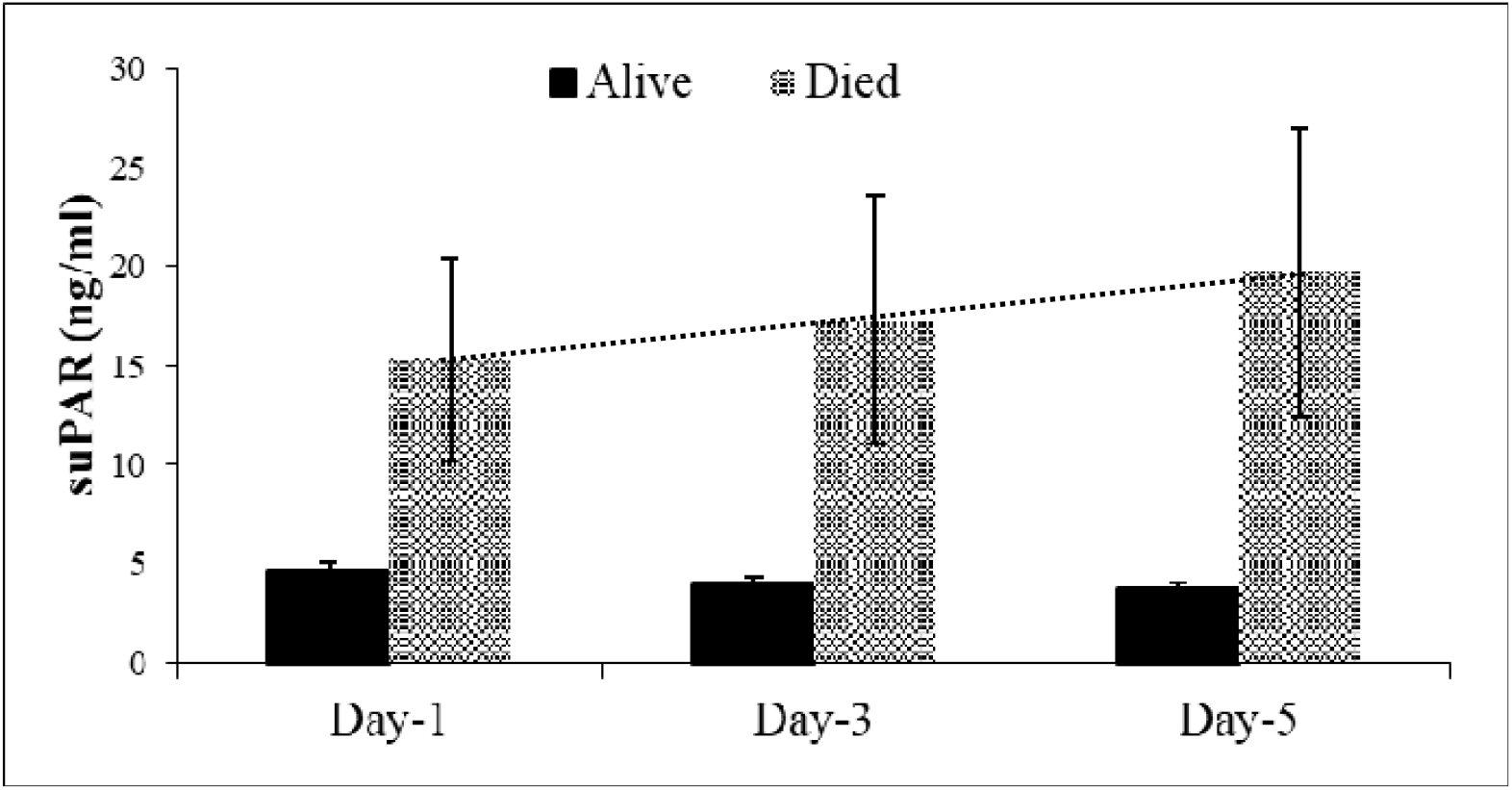
Trend of suPAR in alive and deceased patients. Black Column depicts level of soluble urokinase plasminogen activator receptor (suPAR) on day 1 (4.51 ± 0.46), day 3 (3.95 ± 0.42) and day 5 (3.7 ± 0.34) among live patients (p=0.66) and checked-box column represent the levels in deceased patients on day 1 (15.28 ± 5.12) (p <0.016), day 3 (17.3 ± 6.29) (p <0.011) and day 5 (19.6 ± 7.27) (p < 0.006).

### 3.3 Effect of cormorbidities on levels of suPAR and CRP

Next, the patients were sub-categorized based on their pre-existing chronic co-morbidities. Table 3 shows the levels of suPAR in patients with co-morbidities. Significant increased levels were observed in those patients who had underlying CKD compared to those without CKD as estimated on each day of evaluation [day-1(10.7± 1.3) vs (4.69± 0.83); p <0.001; day-3 (9.9± 1.9) vs (4.3± 0.99); p <0.001; day-5 (8.8± 2.86) vs (4.6± 1.14); p < 0.008].

**Table 3.** Level of suPAR in COVID-19 patients (with/without CKD, with/without HTN, with/without DM)

| Mean ± SEM suPAR(ng/ml) |  |  |  |  |
| --- | --- | --- | --- | --- |
|  | Day-1 | Day-3 | Day-5 | p- value |
| Without CKD (26) | 4.69± 0.83 | 4.3± 0.99 | 4.6± 1.14 | 0.223 |
| With CKD (5) | 10.7± 1.3 | 9.9± 1.9 | 8.8± 2.86 | 0.247 |
| *p-value* | 0.001 | 0.001 | 0.008 |  |
| Without HTN (14) | 3.63 ± 0.38 | 3.19 ± 0.37 | 3.30 ± 0.44 | 0.22 |
| With HTN (17) | 7.13 ± 1.38 | 6.93 ± 1.62 | 6.92 ± 1.87 | 0.66 |
| *p-value* | 0.032 | 0.021 | 0.026 |  |
| Without Diabetes (20) | 4.42 ± 0.53 | 3.92 ± 0.51 | 3.71 ± 0.45 | 0.086 |
| With Diabetes (11) | 7.62 ± 2.03 | 7.64 ± 2.43 | 8.14 ± 2.81 | 0.307 |
| *p-value* | 0.127 | 0.127 | 0.091 |  |

*Number of patients mentioned in parenthesis*
*\*p≤ 0.05 denotes significance and p> 0.05 denotes non-significance*
*Wilcoxon signed rank test P < 0.001 signifies statistical significance of suPAR level between Day 1 and Day 5 among alive patients*

Similarly, COVID-19 patients with pre-existing HTN had significantly increased lev-els of serum suPAR [day-1 (7.13 ± 1.38) vs (3.63 ± 0.38); p<0.032; day-3 (6.93 ± 1.62) vs (3.19± 0.37); p<0.021; day-5 (6.92 ± 1.87) vs (3.30 ± 0.44); p<0.026] than those who did not have underlying HTN (Table 3). However, levels of suPAR did not show statistical risk significance to identify patients with/without diabetes (DM).

Wilcoxon signed rank test was conducted to analysis the significance of CRP level in COVID-19 patients with/without CKD, HTN and DM. Table 4 distinctly showed that CRP levels did not correlate with patient’s pre-existing comorbid conditions except in those patients who had underlying CKD. The levels were significantly elevated [day1 - 122± 33.04 and 63.14± 17.85 respectively; p< 0.03; day3 - 93.6±19.12 and 24.21± 7.82; p< 0.011].

**Table 4.** Comparison of CRP levels in COVID-19patients (with CKD/without CKD, with HTN/without HTN, with DM/without DM)

| Mean ± SEM CRP (mg/dL) |  |  |  |  |
| --- | --- | --- | --- | --- |
|  | Day-1 | Day-3 | Day-5 | p-value |
| with CKD(n=5/31) | 122± 33.04 | 93.6±19.12 | 37.4± 24.7 | **0.247** |
| without CKD(n=14/31) | 63.14± 17.85 | 29.12± 7.17 | 24.21±7.82 | **0.121** |
| **p-value** | **0.03** | **0.011** | **0.513** |  |
| Without HTN (n=6) | 68.16 ± 37.45 | 27.29 ± 11.03 | 20.33±14.21 | **0.607** |
| With HTN (n=13) | 83.46 ±17.84 | 54.76 ± 12.71 | 31.07±10.58 | **0.041** |
| **p-value** | **0.236** | **0.236** | **0.132** |  |
| Without DM (n=12) | 79.42 ± 25.17 | 43.14± 13.7 | 25.08±8.86 | **0.241** |
| With DM (n=7) | 77.2 ± 14.69 | 51.14 ± 12.8 | 32.14±17.85 | **0.156** |
| **p-value** | **0.398** | **0.29** | **0.522** |  |

Number of patients mentioned in parenthesis
\* $p \leq 0.05$ denotes significance; \*\* $p \leq 0.005$ denotes highly significant; $p > 0.05$ denotes non-significance; Wilcoxon signed rank test

Those patients whose hospital stay was ≤ 10 days were significantly (p=0.038) associated with decreased suPAR levels (4.566 ± 0.802) as calculated by Friedman Test (Table 5). An increased trend in the levels of suPAR was observed among those patients requiring more than 10 days of hospital stay.

**Table 5.** Comparison of suPAR levels in COVID-19 patients with respect to Hospital stay.

| SuPAR (mg/dL) |  |  |  |  |
| --- | --- | --- | --- | --- |
| Mean ± SEM | Day-1 | Day-3 | Day-5 | p-value |
| $\leq 10$ Days | $4.566 \pm 0.802$ | $3.714 \pm 0.68$ | $3.508 \pm 0.537$ | **0.038** |
| 10-20 Days | $5.596 \pm 1.125$ | $5.816 \pm 1.259$ | $5.822 \pm 1.462$ | **0.529** |
| $\geq 20$ Days | $8.446 \pm 3.907$ | $8.581 \pm 4.928$ | $9.467 \pm 5.744$ | **0.549** |

Correlation of CRP levels with mortality are shown in Figure 3. Patients who were alive until day-5 showed significantly decreasing levels of CRP (p=0.008), but a statistical non-significant increase was observed among deceased patients (p=0.717).

**Figure 3.**
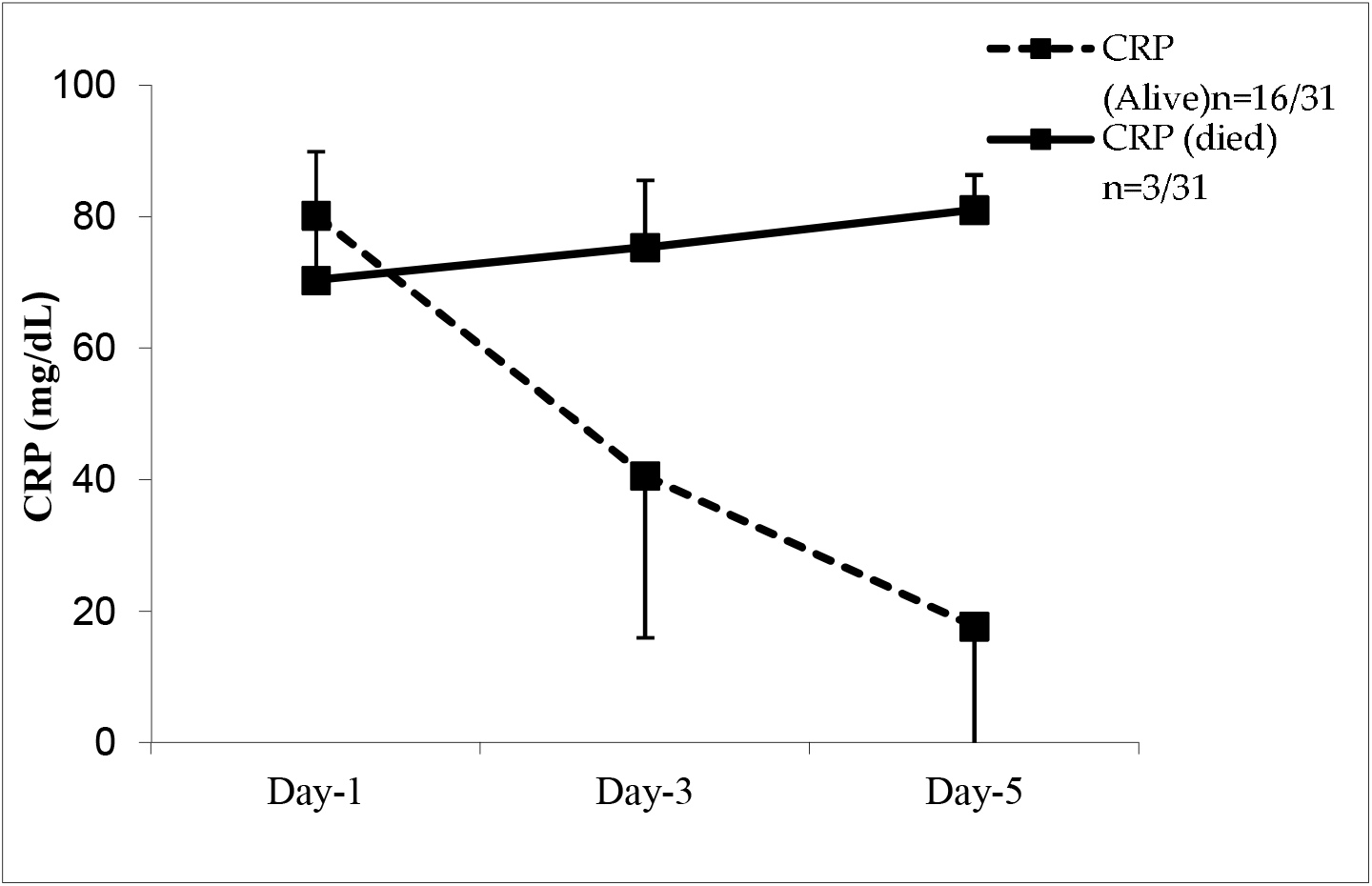
Trend of CRP in alive (n=16/31) and deceased patients (n=3/31), Significant decrease in CRP levels in alive patients from Day-1 to Day-5 (dotted line; p =0.008); No significant difference in the levels of CRP among the deceased patients from Day 1 to Day-5 (straight line; p=0.717).

## 4. Discussion

In the present study, 31 consecutive hospitalized COVID-19 patients observed a male dominance with 80% of the patients more than 50 years of age. Apart from age factor, inflammatory status during SARS-CoV-2 infection have been cited to play role in disease severity. For identification of this, suPAR levels have shown to be sensitive biomarker in patients with severe COVID-19. In earlier studies conducted at our center on non-COVID patients, suPAR had helped in differentiation between survivors and/or non-survivors as well prognosticated critically-ill patients in intensive care and thus, concluding that it can predict/prognosticate the severity [13,14].

Our data showed higher suPAR levels (ranged from 15.28 ± 5.12 on day 1 to 19.6 ± 7.27 on day 5) were found to be associated with the risk of mortality. Elevated levels of suPAR represents strong immune cell activation [15,16,17] and has shown to be correlative in critically ill patients with sepsis and systemic inflammation response syndrome.

In order to find a sensitive biomarker that can stratify severe COVID-19 patients admitted with moderate disease status, we evaluated CRP serially similar to suPAR estimation. The data from our study did not observe a significant correlation. Our result is consistent with the study by Hashem et al, 2021, wherein the authors reported that CRP was not a predictor for severity [18]. CRP has low sensitivity and unable to distinguish be-tween viral and bacterial sepsis [19]. A recent pre-print by Ji W and team, reports that Se-rum Amyloid A was a better prognostic marker than CRP during SARS-Cov-2 infection.

More recently, Dholariya S et al, 2021 reviewed the literature to identify biomarkers that can risk stratify and prognostify SARS-Cov-2 cases [20]. The authors suggested P-SEP (presepsin), s-TREM-1 (soluble form of the triggering receptor expressed on myeloid cells) and suPAR as novel biomarkers that positively correlated with infection severity.

We observed that all those patients who died had progressed to severe COVID and developed ARDS. Chen D et al, 2019 observed serum suPAR levels predicted increased risk of ARDS in patients with sepsis [21]. Furthermore, its upregulation was observed to be directly proportionate with higher disease severity, inflammation and increased mortality in patients with sepsis and ARDS [22]. Rovina N et al (2020) suggesting suPAR as an ear-ly predictor of severe respiratory failure in patients with COVID-19 pneumonia [23].

Chudasama YV et al (2021) suggested that the multi-morbidity index encompassing hypertension, diabetes mellitus and chronic kidney disease could help in identifying se-verity of COVID-19 [24]. A study by Gansevoor et al (2020) and team found that patients with severe CKD had increased risk of mortality due to COVID 19, which was greater than other known high-risk groups (hypertension and diabetes) [25]. In our study, we evaluated the levels of suPAR and CRP in the sub-categorized COVID-19 patients, based on their pre-existing comorbities (CKD, HTN, and DM). Levels of suPAR were significantly in-creased in those patients who had underlying CKD and HTN compared to those without these conditions. Our data corroborated with findings by Napolitano F et al wherein the authors suggest CKD as a significant risk factor for severity and mortality among ICU COVID-19 Patients [26]. Earlier studies have evaluated the role of suPAR in chronic dis-eases conditions, coronary artery disease severity [27], cardiovascular disease in chronic kidney disease [28] and others like diabetes, cancer and liver disease [22]. Azamet et al, 2020 suggested suPAR may be associated with pathophysiology of AKI in COVID-19 patients [29]. Very recently, a clinical trial [NCT04590794] has been initiated to determine the predictive utility of suPAR level measured at admission for risk stratification and mortality among COVID-19 patients.

Another observation in our study was that longer the patients were required to stay in the hospital due to their criticality, suPAR level were found to be increase proportionally. Earlier studies have reported the length of hospital stay as a measure of disease severity [30,31].

During the period of our study, Delta variant had emerged and peaked in the month of March, though, earlier variant were yet prevalent. Hence, presence of these diverse variants in our study population can affected the outcome of the data. The patients enrolled here were not a part of any study involving sequencing. Another technical limitation was that the ELISA kit used in the study though can detect antibodies against human uPAR1 isoform (full length and DIIDIII fragment), most of the alternate isoforms of human uPAR can go undetected. Different isoforms of suPAR may be induced by the diverse variants of SARS-CoV-2. Therefore, we intend further studying the presence of different uPAR isoforms, which will be able to shed important insights in COVID-19 pathogenesis.

## 5. Conclusions

This study was conducted at the beginning of second wave which observed a high mortality rate and a very rapid deterioration in the general condition of patients, leaving many of them debilitated for longer period. As per the WHO, the disease is here to stay, the reason being various epigenetic factors favoring its existence which are leading to multiple mutations in the virus genome. Our data introduces a new aid available to clinicians for better assessment and timely management of moderate COVID-19 patients. Levels of suPAR were found to be directly related to mortality rates in the patients. Additionally, existing co-morbid conditions - chronic kidney diseases and hypertension can influence the level of suPAR in COVID-19, independent of age factor. Thus suggesting that circulating suPAR levels are a sensitive prognostic biomarker in COVID-19 patients. 6. Patents

## Data Availability

All data produced in the present work are contained in the manuscript

## Author Contributions

The manuscript was written through contributions of all authors. All authors have given approval to the final version of the manuscript. AK enrolled the patients, aided in data analysis and advised editing the manuscript. PR collated the data, analyzed it and framed the manuscript. TB contributed in clinical data acquisition and its analysis. RH performed the in-vitro suPAR assays. SC conceptualized, guided, framed and edited the manuscript.

## Institutional Review Board Statement

The ethics committee of Sir Ganga ram Hospital (#EC/08/20/1711) approved the study.

## Informed Consent Statement

Informed consent obtained from all subjects involved in the study for their voluntary participation and consent for publication.

## Acknowledgments

The authors would like to thank Mr. Devinder Soni from ViroGates for providing us with the kits to perform suPAR and Mr. Jasper for his advices. They both had no role in planning, conduct and analysis of this study. The authors would like to acknowledge Parul Chugh, Biostatistician, Sir Ganga Ram Hospital, New Delhi for statistical analysis. A heartfelt thanks to the enrolled COVID-19 patients and their families for co-operating with the clinical and the research team.

## References

1. Worldometer – Real time world statistics.

2. http://www.worldometers.info/coronavirus/country/india/

3. Ali N. Elevated level of C-reactive protein may be an early marker to predict risk for severity of COVID-19. J Med Virol. 2020 Nov;92(11):2409–2411. doi: 10.1002/jmv.26097. Epub 2020 Jun 9. PMID: 32516845; PMCID: PMC7301027.

4. Wang G, Wu C, Zhang Q, Wu F, Yu B, Lv J, Li Y, Li T, Zhang S, Wu C, Wu G, Zhong Y. C-Reactive Protein Level May Predict the Risk of COVID-19 Aggravation. Open Forum Infect Dis. 2020 Apr 29;7(5):ofaa153. doi: 10.1093/ofid/ofaa153. PMID: 32455147; PMCID: PMC7197542.

5. Tan C, Huang Y, Shi F, Tan K, Ma Q, Chen Y, Jiang X, Li X. C-reactive protein correlates with computed tomographic findings and predicts severe COVID-19 early. J Med Virol. 2020 Jul;92(7):856–862. doi: 10.1002/jmv.25871. Epub 2020 Apr 25. PMID: 32281668; PMCID: PMC7262341.

6. Pepys MB, Hirschfield GM. C-reactive protein: a critical update. J Clin Invest. 2003 Jun;111(12):1805–12. doi: 10.1172/JCI18921. Erratum in: J Clin Invest. 2003 Jul;112(2):299. PMID: 12813013; PMCID: PMC161431.

7. Sproston NR, Ashworth JJ. Role of C-Reactive Protein at Sites of Inflammation and Infection. Front Immunol. 2018 Apr 13;9:754. doi: 10.3389/fimmu.2018.00754. PMID: 29706967; PMCID: PMC5908901..

8. Volanakis JE. Human C-reactive protein: expression, structure, and function. Mol Immunol. 2001 Aug;38(2-3):189–97. doi: 10.1016/s0161-5890(01)00042-6. PMID: 11532280.

9. Montuori N, Ragno P. Multiple activities of a multifaceted receptor: roles of cleaved and soluble uPAR. Front Biosci (Landmark Ed). 2009 Jan 1;14(7):2494–503. doi: 10.2741/3392. PMID: 19273214.

10. Wittenhagen P, Kronborg G, Weis N, Nielsen H, Obel N, Pedersen SS, Eugen-Olsen J. The plasma level of soluble urokinase receptor is elevated in patients with Streptococcus pneumoniae bacteraemia and predicts mortality. Clin Microbiol Infect. 2004 May;10(5):409–15. doi: 10.1111/j.1469-0691.2004.00850.x. PMID: 15113317.

11. Rasmussen LJ, Ladelund S, Haupt TH, Ellekilde G, Poulsen JH, Iversen K, Eugen-Olsen J, Andersen O. Soluble urokinase plasminogen activator receptor (suPAR) in acute care: a strong marker of disease presence and severity, readmission and mortality. A retrospective cohort study. Emerg Med J. 2016 Nov;33(11):769–775. doi: 10.1136/emermed-2015-205444. Epub 2016 Sep 2. PMID: 27590986; PMCID: PMC5136705.

12. Eugen-Olsen J. suPAR - a future risk marker in bacteremia. J Intern Med. 2011 Jul;270(1):29–31. doi: 10.1111/j.1365-2796.2011.02372.x. Epub 2011 Mar 24. PMID: 21366732.

13. covid19treatmentgudelines.nih.gov/overview/clinical-spectrum/

14. Sharma A, Ray S, Mamidipalli R, Kakar A, Chugh P, Jain R, Ghalaut MS, Choudhury S. A Comparative Study of the Diagnostic and Prognostic Utility of Soluble Urokinase-type Plasminogen Activator Receptor and Procalcitonin in Patients with Sepsis and Systemic Inflammation Response Syndrome. Indian J Crit Care Med. 2020 Apr;24(4):245–251. doi: 10.5005/jp-journals-10071-23385. PMID: 32565634; PMCID: PMC7297249.

15. Kumar P, Kakar A, Gogia A, Waziri N. Evaluation of soluble urokinase-type plasminogen activator receptor (suPAR) quick test for triage in the emergency department. J Family Med Prim Care. 2019 Dec 10;8(12):3871–3875. doi: 10.4103/jfmpc.jfmpc_116_19. PMID: 31879628; PMCID: PMC6924241.

16. Yilmaz G, Köksal I, Karahan SC, Mentese A. The diagnostic and prognostic significance of soluble urokinase plasminogen activator receptor in systemic inflammatory response syndrome. Clin Biochem. 2011 Oct;44(14-15):1227–30. doi: 10.1016/j.clinbiochem.2011.07.006. Epub 2011 Jul 26. PMID: 21816136..

17. Enocsson H, Wirestam L, Dahle C, Padyukov L, Jönsen A, Urowitz MB, Gladman DD, Romero-Diaz J, Bae SC, Fortin PR, Sanchez-Guerrero J, Clarke AE, Bernatsky S, Gordon C, Hanly JG, Wallace DJ, Isenberg DA, Rahman A, Merrill JT, Ginzler E, Alarcón GS, Chatham WW, Petri M, Khamashta M, Aranow C, Mackay M, Dooley MA, Manzi S, Ramsey-Goldman R, Nived O, Steinsson K, Zoma AA, Ruiz-Irastorza G, Lim SS, Kalunian KC, Inanc M, van Vollenhoven RF, Ramos-Casals M, Kamen DL, Jacobsen S, Peschken CA, Askanase A, Stoll T, Bruce IN, Wetterö J, Sjöwall C. Soluble urokinase plasminogen activator receptor (suPAR) levels predict damage accrual in patients with recent-onset systemic lupus erythematosus. J Autoimmun. 2020 Jan;106:102340. doi: 10.1016/j.jaut.2019.102340. Epub 2019 Oct 17. PMID: 31629628.

18. Hamie L, Daoud G, Nemer G, Nammour T, El Chediak A, Uthman IW, Kibbi AG, Eid A, Kurban M. SuPAR, an emerging biomarker in kidney and inflammatory diseases. Postgrad Med J. 2018 Sep;94(1115):517–524. doi: 10.1136/postgradmedj-2018-135839. Epub 2018 Sep 3. PMID: 30177549.

19. Hashem, M.K., Khedr, E.M., Daef, E., Mohamed-Hussein, A., Mostafa, E.F., Hassany, S.M., Galal, H., Hassan, S.A., Galal, I., Amin, M.T. and Hassan, H.M.,Prognostic biomarkers in COVID-19 infection: value of anemia, neutrophil-to-lymphocyte ratio, platelet-to-lymphocyte ratio, and D-dimer. The Egyptian Journal of Bronchology 2021, 15(1), (pp.1–9).doi.org

20. Ji, W., Bishnu, G., Cai, Z. and Shen, X., Analysis clinical features of COVID-19 infection in secondary epidemic area and report potential biomarkers in evaluation. MedRxiv.2020,doi.org/10.1101/2020.03.10.20033613.

21. Dholariya S, Parchwani DN, Singh R, Radadiya M, Katoch CDS. Utility of P-SEP, sTREM-1 and suPAR as Novel Sepsis Biomarkers in SARS-CoV-2 Infection. Indian J Clin Biochem. 2021 Oct 6:1–8. doi: 10.1007/s12291-021-01008-6. Epub ahead of print. PMID: 34642555; PMCID: PMC8494168.

22. Chen D, Wu X, Yang J, Yu L. Serum plasminogen activator urokinase receptor predicts elevated risk of acute respiratory distress syndrome in patients with sepsis and is positively associated with disease severity, inflammation and mortality. Exp Ther Med. 2019 Oct;18(4):2984–2992. doi: 10.3892/etm.2019.7931. Epub 2019 Aug 20. PMID: 31555383; PMCID: PMC6755407.

23. Haupt TH, Petersen J, Ellekilde G, Klausen HH, Thorball CW, Eugen-Olsen J, Andersen O. Plasma suPAR levels are associated with mortality, admission time, and Charlson Comorbidity Index in the acutely admitted medical patient: a prospective observational study. Crit Care. 2012 Jul 23;16(4):R130. doi: 10.1186/cc11434. PMID: 22824423; PMCID: PMC3580714.

24. Rovina N, Akinosoglou K, Eugen-Olsen J, Hayek S, Reiser J, Giamarellos-Bourboulis EJ. Soluble urokinase plasminogen activator receptor (suPAR) as an early predictor of severe respiratory failure in patients with COVID-19 pneumonia. Crit Care. 2020 Apr 30;24(1):187. doi: 10.1186/s13054-020-02897-4. PMID: 32354367; PMCID: PMC7191969.

25. Chudasama YV, Zaccardi F, Gillies CL, Razieh C, Yates T, Kloecker DE, Rowlands AV, Davies MJ, Islam N, Seidu S, Forouhi NG, Khunti K. Patterns of multimorbidity and risk of severe SARS-CoV-2 infection: an observational study in the U.K. BMC Infect Dis. 2021 Sep 4;21(1):908. doi: 10.1186/s12879-021-06600-y. PMID: 34481456; PMCID: PMC8418288.

26. Gansevoort RT, Hilbrands LB. CKD is a key risk factor for COVID-19 mortality. Nat Rev Nephrol. 2020 Dec;16(12):705–706. doi: 10.1038/s41581-020-00349-4. PMID: 32848205; PMCID: PMC7447963.

27. Napolitano F, Di Spigna G, Vargas M, Iacovazzo C, Pinchera B, Spalletti Cernia D, Ricciardone M, Covelli B, Servillo G, Gentile I, Postiglione L, Montuori N. Soluble Urokinase Receptor as a Promising Marker for Early Prediction of Outcome in COVID-19 Hospitalized Patients. J Clin Med. 2021 Oct 24;10(21):4914. doi: 10.3390/jcm10214914. PMID: 34768433; PMCID: PMC8584815.

28. Eapen DJ, Manocha P, Ghasemzadeh N, Patel RS, Al Kassem H, Hammadah M, Veledar E, Le NA, Pielak T, Thorball CW, Velegraki A, Kremastinos DT, Lerakis S, Sperling L, Quyyumi AA. Soluble urokinase plasminogen activator receptor level is an independent predictor of the presence and severity of coronary artery disease and of future adverse events. J Am Heart Assoc. 2014 Oct 23;3(5):e001118. doi: 10.1161/JAHA.114.001118. Erratum in: J Am Heart Assoc. 2015 Jan;4(1):e000563. Ghasemzedah, Nima [Corrected to Ghasemzadeh, Nima]. Erratum in: J Am Heart Assoc. 2015 Jan 5;4(1):e000563. PMID: 25341887; PMCID: PMC4323820.

29. Meijers B, Poesen R, Claes K, Dietrich R, Bammens B, Sprangers B, Naesens M, Storr M, Kuypers D, Evenepoel P. Soluble urokinase receptor is a biomarker of cardiovascular disease in chronic kidney disease. Kidney Int. 2015 Jan;87(1):210–6. doi: 10.1038/ki.2014.197. Epub 2014 Jun 4. PMID: 24897037.

30. Azam TU, Shadid HR, Blakely P, O ‘Hayer P, Berlin H, Pan M, Zhao P, Zhao L, Pennathur S, Pop-Busui R, Altintas I, Tingleff J, Stauning MA, Andersen O, Adami ME, Solomonidi N, Tsilika M, Tober-Lau P, Arnaoutoglou E, Keitel V, Tacke F, Chalkias A, Loosen SH, Giamarellos-Bourboulis EJ, Eugen-Olsen J, Reiser J, Hayek SS; International Study of Inflammation in COVID-19. Soluble Urokinase Receptor (SuPAR) in COVID-19-Related AKI. J Am Soc Nephrol. 2020 Nov;31(11):2725–2735. doi: 10.1681/ASN.2020060829. Epub 2020 Sep 22. PMID: 32963090; PMCID: PMC7608953.

31. Reisinger AC, Niedrist T, Posch F, Hatzl S, Hackl G, Prattes J, Schilcher G, Meißl AM, Raggam RB, Herrmann M, Eller P. Soluble urokinase plasminogen activator receptor (suPAR) predicts critical illness and kidney failure in patients admitted to the intensive care unit. Sci Rep. 2021 Sep 1;11(1):17476. doi: 10.1038/s41598-021-96352-1. PMID: 34471146; PMCID: PMC8410930.

32. Rees EM, Nightingale ES, Jafari Y, Waterlow NR, Clifford S B Pearson CA, Group CW, Jombart T, Procter SR, Knight GM. COVID-19 length of hospital stay: a systematic review and data synthesis. BMC Med. 2020 Sep 3;18(1):270. doi: 10.1186/s12916-020-01726-3. PMID: 32878619; PMCID: PMC7467845.

